# Internalizing Psychiatric Symptoms in People with Mosaicism for Trisomy 21

**DOI:** 10.1101/2024.06.19.24309168

**Authors:** Ruth C. Brown, Allison D’Aguilar, Quinn Hurshman, Rebekah NailorZee, Timothy P. York, George Capone, Ananda B. Amstadter, Colleen Jackson-Cook

## Abstract

People with mosaicism for trisomy 21 have been shown to exhibit the many of same phenotypic traits present in people with non-mosaic Down syndrome, but with varying symptom severity. However, the behavioral phenotype of people with mosaic Down syndrome (mDS) has not been well characterized. This study aimed to examine the prevalence of self-report and caregiver-report symptoms of depression and anxiety among a sample of 62 participants with mDS aged 12 - 46, and assess their association with the percentage of trisomy 21 in blood and/or buccal mucosa cells. The results showed that 53% of the participants reported clinically significant depression symptoms and 76% reported clinically significant anxiety symptoms. No clear associations were observed between the percentage of trisomic cells and total anxiety or depression, but a significant positive association between the proband-reported specific fears subscale and the percentage of trisomic cells in buccal specimens was detected (*r* = .43, *p* = .007). This study highlights the high occurrence of depression and anxiety symptoms in individuals with mDS and the need for routine assessment to optimize their care. It also demonstrates the ability of people with mDS to complete these evaluations, thereby supporting their inclusion in research studies/clinical trials.

---

Down syndrome (DS), which results from a trisomic imbalance for chromosome 21, is the most common known genetic cause of intellectual disability (ID) (Canfield et al., 2014), affecting approximately 1 in 600 live births in the United States (Parker et al., 2010). Mosaicism for a trisomic imbalance of chromosome 21 (mDS) occurs in approximately 2%-4% of people diagnosed with DS (Papavassiliou et al., 2015). People with mDS typically have two cell lines that originated from a single zygote, including cells with a trisomic imbalance for chromosome 21 and cells with a disomic chromosomal complement (Nussbaum et al., 2001).

Researchers have noted a positive correlation between the percentage of trisomic cells in persons with mDS and the presentation of phenotypic characteristics (Papavassiliou et al., 2015). Furthermore, the observed karyotype-phenotype correlation values varied between tissues, possibly reflecting (at least in part), the embryonic developmental origins of the tissue/trait. For example, the percentage of trisomy in lymphocytes (mesodermal origin) was more highly correlated with risk for congenital heart defects (heart tissue is also mesodermal in origin) and the percentage of trisomy in buccal mucosa cells (ectodermal origin) was more highly correlated with IQ (brain/neural tissue is ectodermal in origin) (Papavassiliou et al., 2009; Winning & Townsend, 2000).

There is evidence that people with non-mosaic DS are more prone to depression and anxiety than the general population, as well as people with other forms of ID (Dykens, 2007; Walker et al., 2011). Indeed, anxiety and depression have been considered as potential “behavioral phenotypes” of people with DS, implying a potential direct or indirect link with the trisomy 21 karyotype (G. Capone et al., 2006; Dykens, 2007; Levitas et al., 2018). However, to our knowledge, the prevalence of internalizing disorders among people with mDS have not yet been reported, nor has the extent to which the severity of internalizing symptoms may be associated with “dosage imbalance” (e.g. percent trisomy). To inform best practices in patient care and treatment planning, research that sheds light on the prevalence of internalizing psychiatric symptoms, such as depression and anxiety, in people with mDS has been undertaken.

This study aimed to characterize symptoms of depression and anxiety in adolescents and adults with mDS using both self-report and caregiver-report measures. We also examined potential relationships between blood- and buccal mucosa cell-derived trisomy 21 percentages with symptoms of depression and anxiety to test the hypothesis that the presentation of behavioral traits is positively correlated with the percentage of cells having a trisomic imbalance.

## Methods

### Participants and Procedures

All study procedures were reviewed and approved by the Virginia Commonwealth University Institutional Review Board (HM20007318, HM15281). Youth and adult participants with mDS (probands) and their parents were recruited through multiple sources, including annual research retreats of the International Mosaic Down Syndrome Association (IMDSA) via a flyer and study description listed in conference materials, previous study participants who indicated interest in participating in new projects (i.e., previous participants in research lead by CJC), outreach to DS-focused parent advocacy organizations around the US, and through the NIH DS-Connect registry (Peprah et al., 2015).

Recruitment occurred from July 2013 to July 2021. Consent was obtained from adult participants and/or parent participants. Minor participants and adults under the legal guardianship of their parent were asked for assent. An interviewer met with proband participants face-to-face and was present throughout testing to assist with measure completion in the form of reading questions and response options to participants. Parents completed assessments independently.

In March 2020, the COVID-19 pandemic interrupted recruitment and data collection, which was then shifted to online methods. For online participants, research staff met with parents and probands via Zoom to conduct consent/assent procedures and scheduled one or more virtual data collection appointments. Questionnaires were converted to online surveys using Research Electronic Data Capture (REDCap (Harris et al., 2009)). Trained research staff met with probands via Zoom and used screen-sharing to display the survey questions, read them aloud to the participant, and record the participant’s responses. Parents were allowed to be in the room and support the proband with technology or facilitating communication, but were asked to refrain from influencing the proband’s response. Parents completed their surveys via REDCap independently.

## Materials

### Demographics

Guided by a demographic form completed by adult probands and parents, information was collected on age, gender, DS diagnosis (i.e., mosaic Down syndrome, Trisomy 21, other), educational and occupational status and history of diagnosis or treatment for depression, anxiety, or other behavioral problems. Parents were also asked if they currently had concerns about their offspring’s depression, anxiety, or other behavioral problems. Parents also provided an estimate of their offspring’s level of cognitive impairment using IQ bands (e.g. severe intellectual disability (IQ < 49), moderate intellectual disability (IQ = 50-69), mild intellectual disability (IQ = 70-79), average (IQ = 80-109), above average (IQ > 110)). IQ bands were coded such that higher scores reflected higher estimated IQ. In instances where both the parent and proband provided demographic data, the parent data was utilized.

### Percentage of trisomic cells

Participants had the option to provide a biological specimen(s) and/or medical records to confirm the percentage of trisomic cells. The submission of biological specimens was limited, due to COVID-19 restrictions on face-to-face meetings or research specimen procurement. However, for the earlier study participants, the probands could elect to provide a blood specimen and/or a buccal smear specimen. The percentage of trisomic cells present in these specimens was determined, using fluorescence in situ hybridization (FISH) technology. For the peripheral blood specimens, 1000 interphase nuclei were scored; for the buccal mucosa specimens, 500 interphase nuclei were scored, as previously described (Papavassiliou et al., 2009). A subset of participants provided copies of proband diagnostic reports and/or consent for this information to be extracted from their medical record, rather than biological specimens. The diagnostic reports were typically completed in the neonatal period. In the event that participants had multiple measures available, the earliest available recorded value was used.

### Glasgow Depression Scale for People with a Learning Disability

(GDS-LD; Cuthill et al., 2003). The GDS-LD, a self-report measure developed for people with ID, was completed by probands. It contains 20 items based on DSM-IV and ICD-10 criteria for depression. Items are rated on a 3-point scale (0 = never/no, 1 = sometimes, 2 = always/a lot) to reflect how they have “been feeling lately”. Possible scores can range from 0–40, with higher scores reflecting greater depression symptoms. A cut-off score of equal to or greater than 13 has demonstrated a sensitivity of 96% and specificity of 90% to distinguish people with mild to moderate ID with and without depression (Cuthill et al., 2003). Cronbach’s alpha for the proband report GDS-LD in this cohort was = 0.83.

Parents completed the Glasgow Depression Scale–Carer Supplement (GDS-CS; Cuthill et al., 2003). The GDS-CS is a 16-item measure rated on a 3-point scale (0 = never/no, 1 = sometimes, 2 = always/a lot) of symptoms observed in the last week.

Possible scores range from 0 to 32. There is no recommended cut-off score for the GDS-CS. Cronbach’s alpha for the GDS-CS in this cohort was = 0.81

### Glasgow Anxiety Scale for People with Intellectual Disability

(GAS-ID; (Mindham & Espie, 2003)). The GAS-ID is a 27-item self-report measure of anxiety over the past week that was developed for people with ID and was competed by probands.

The GAS-ID includes three scales to assess worry (10 items), specific fears (9 items), and physiological symptoms (8 items) on a 3-point scale (0 = never, 1 = sometimes, 2 = always). Possible scores for the total anxiety scale can range from 0 to 54, with a recommended clinical cut-off score of 13. The internal consistency of the GAS-ID in the study cohort was very good (Cronbach’s *α* = 0.87).

Parents completed the Glasgow Anxiety Scale–Carer Supplement (GAS-CS), a modified version of the GAS-ID that was created for the current study by altering the wording to refer to the probands (e.g., “do you” was changed to “does he/she”). The internal consistency of the GAS-CS in the current sample was very good (Cronbach’s *α* = 0.85). There is no recommended clinical cut-off score for the GAS-CS.

### Data Analysis

Analyses were conducted using the R programming environment (R Core Team, 2022). Means and standard deviations were used to describe the average severity of depression and anxiety symptoms by rater. Pearson correlations were used to estimate bivariate relationships between parent and proband reported symptoms, IQ bands, and percent trisomy in blood or buccal specimens. Multiple regression methods tested for a linear relationship between the percentage of trisomic cells and proband- and parent-reported internalizing symptoms while controlling for the influence of age and gender.

## Results

### Descriptive Statistics

Data on a total of 62 probands were included in the study. This included 9 independent probands who participated without a parent, and 11 parents who completed the parent-report assessments without their proband, either because the proband was unable or uninterested in completing the self-report assessments.

Probands with mDS were aged 12 – 46 years (M = 23.52, SD = 9.68), 63% female, and 92% identified as white, based on self-report or parent-report responses (Table 1). We compared all demographic and outcome variables of interest that were collected pre- and post-pandemic, and found no significant differences, thus, the data was analyzed and presented together. In Table 1, instead of just reporting overall values, you might consider reporting also by quartiles of mDS percentages (or lower/upper 50^th^ percentile)

**Table 1.**
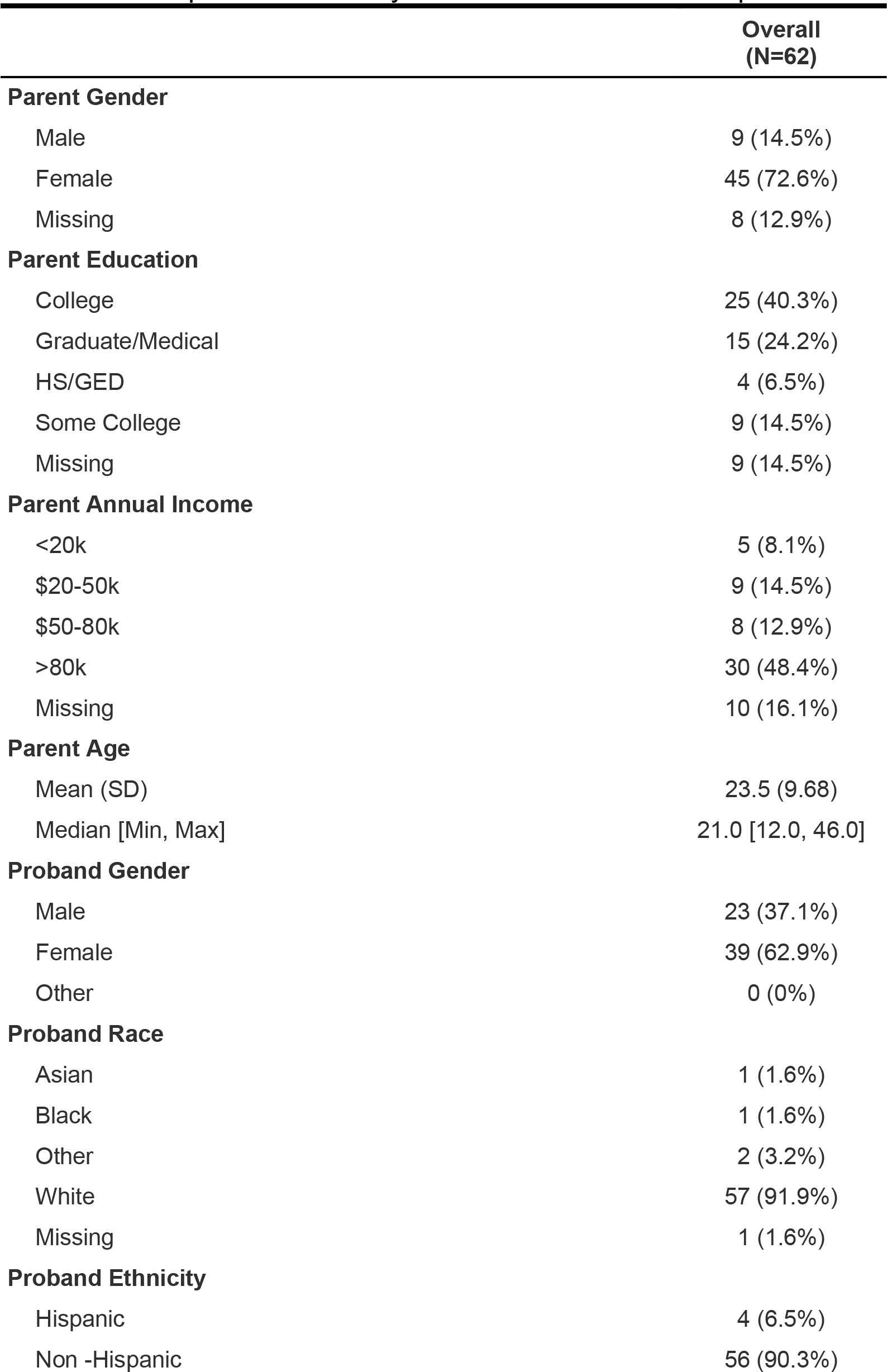
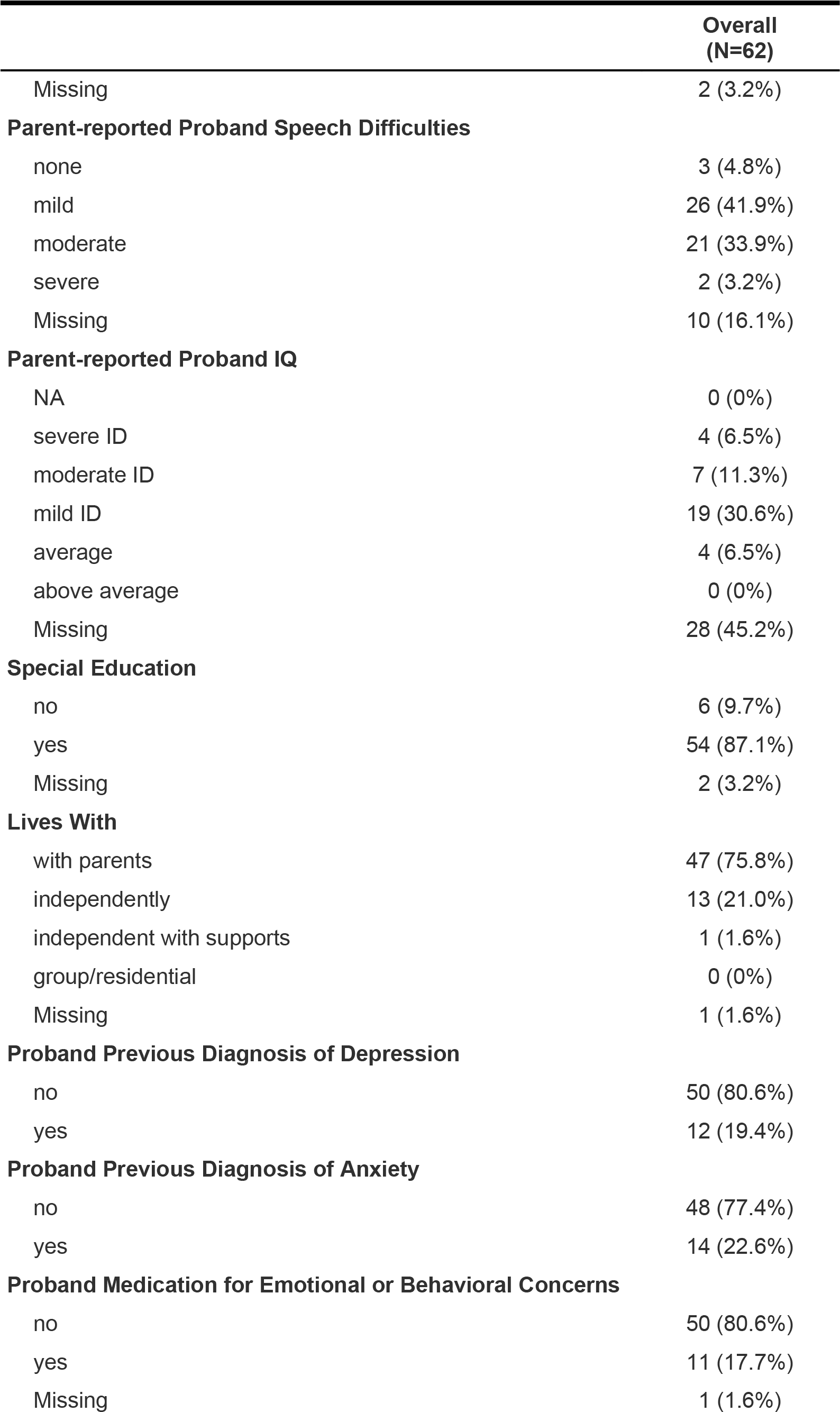
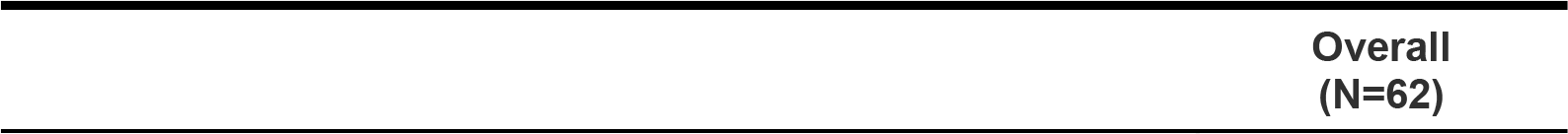
Descriptive Statistics by Parent and/or Proband Report.

|  | Overall<br>(N=62) |
| --- | --- |
| **Parent Gender** |  |
| Male | 9 (14.5%) |
| Female | 45 (72.6%) |
| Missing | 8 (12.9%) |
| **Parent Education** |  |
| College | 25 (40.3%) |
| Graduate/Medical | 15 (24.2%) |
| HS/GED | 4 (6.5%) |
| Some College | 9 (14.5%) |
| Missing | 9 (14.5%) |
| **Parent Annual Income** |  |
| <20k | 5 (8.1%) |
| \$20-50k | 9 (14.5%) |
| \$50-80k | 8 (12.9%) |
| >80k | 30 (48.4%) |
| Missing | 10 (16.1%) |
| **Parent Age** |  |
| Mean (SD) | 23.5 (9.68) |
| Median [Min, Max] | 21.0 [12.0, 46.0] |
| **Proband Gender** |  |
| Male | 23 (37.1%) |
| Female | 39 (62.9%) |
| Other | 0 (0%) |
| **Proband Race** |  |
| Asian | 1 (1.6%) |
| Black | 1 (1.6%) |
| Other | 2 (3.2%) |
| White | 57 (91.9%) |
| Missing | 1 (1.6%) |
| **Proband Ethnicity** |  |
| Hispanic | 4 (6.5%) |
| Non -Hispanic | 56 (90.3%) |
| Missing | 2 (3.2%) |
| **Parent-reported Proband Speech Difficulties** |  |
| none | 3 (4.8%) |
| mild | 26 (41.9%) |
| moderate | 21 (33.9%) |
| severe | 2 (3.2%) |
| Missing | 10 (16.1%) |
| **Parent-reported Proband IQ** |  |
| NA | 0 (0%) |
| severe ID | 4 (6.5%) |
| moderate ID | 7 (11.3%) |
| mild ID | 19 (30.6%) |
| average | 4 (6.5%) |
| above average | 0 (0%) |
| Missing | 28 (45.2%) |
| **Special Education** |  |
| no | 6 (9.7%) |
| yes | 54 (87.1%) |
| Missing | 2 (3.2%) |
| **Lives With** |  |
| with parents | 47 (75.8%) |
| independently | 13 (21.0%) |
| independent with supports | 1 (1.6%) |
| group/residential | 0 (0%) |
| Missing | 1 (1.6%) |
| **Proband Previous Diagnosis of Depression** |  |
| no | 50 (80.6%) |
| yes | 12 (19.4%) |
| **Proband Previous Diagnosis of Anxiety** |  |
| no | 48 (77.4%) |
| yes | 14 (22.6%) |
| **Proband Medication for Emotional or Behavioral Concerns** |  |
| no | 50 (80.6%) |
| yes | 11 (17.7%) |
| Missing | 1 (1.6%) |

|  |
| --- |
| Overall<br>(N=62) |

### Internalizing Symptoms

Scores on the GDS were M = 13.63, SD = 6.22. Using the recommended cutoff score of 13 on the proband version, 26 (53%) reported clinically significant depression symptoms. Scores on the parent-rated GDS-CS were M = 7.55, SD = 6.22. There is no recommended cutoff for the GDS-CS.

Scores on the Total Anxiety subscale of the GAS-ID were M = 19.76, SD = 8.84. Using the recommended cutoff score of 13 on the proband version, 76% reported clinically significant anxiety symptoms. Scores on the parent-rated GAS-CS were M = 13.6, SD = 8.84. There is no recommended cutoff for the GAS-CS. Scores on the Worries, Fears, and Physical Symptoms subscales are found in Table 2.

**Table 2.**
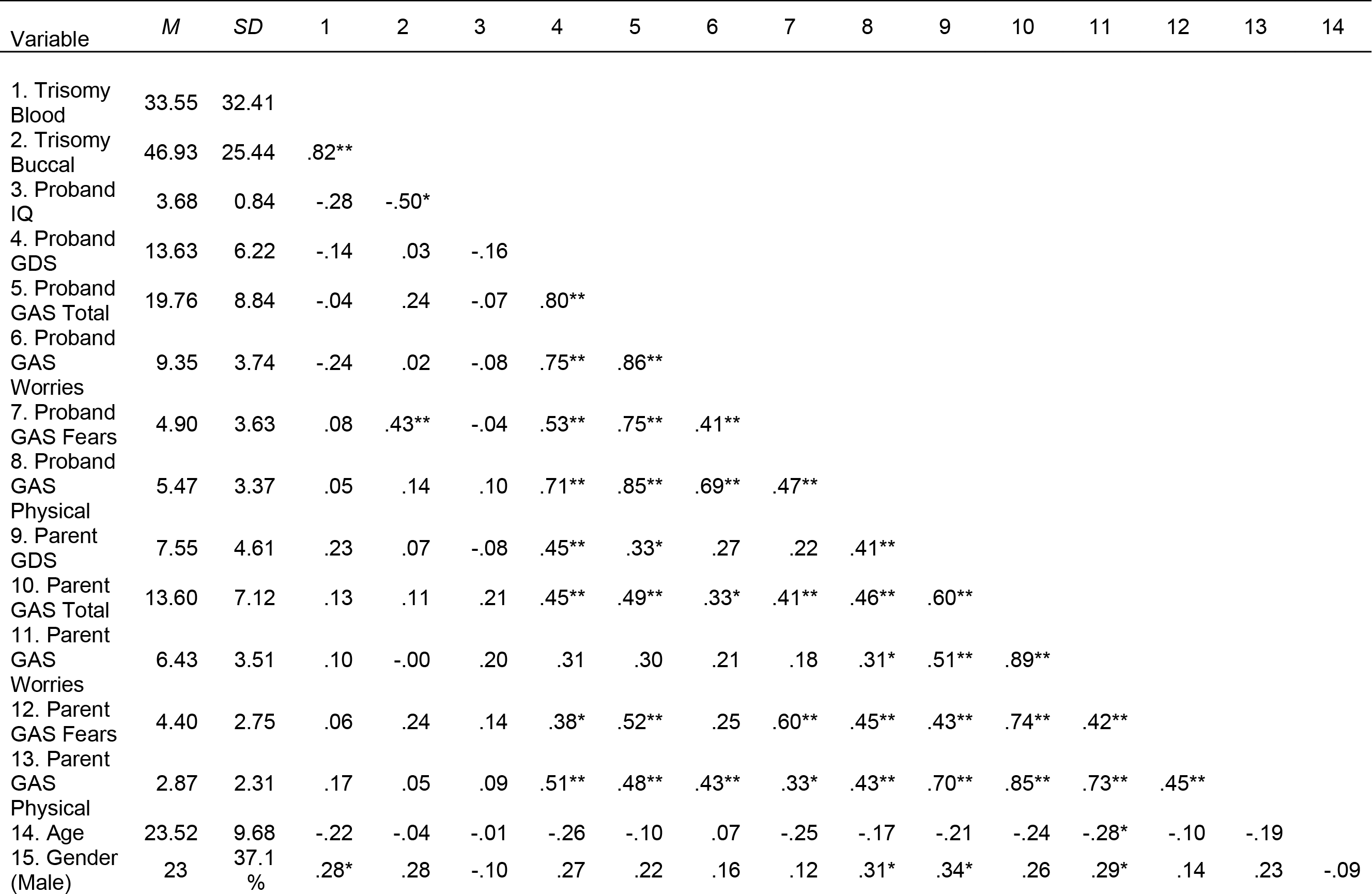
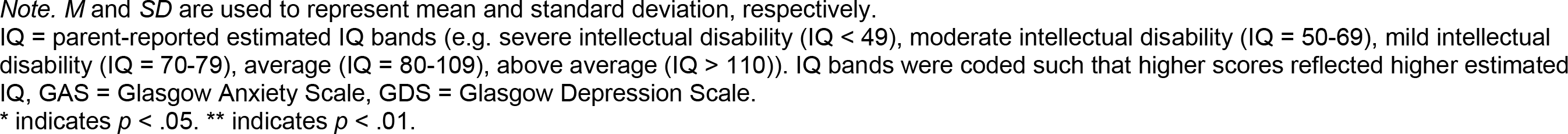
Means, Standard Deviations, and Correlations of Trisomy Percentages with Parent and Proband Reported Internalizing Symptoms.

| Variable | M | SD | 1 | 2 | 3 | 4 | 5 | 6 | 7 | 8 | 9 | 10 | 11 | 12 | 13 | 14 |
| --- | --- | --- | --- | --- | --- | --- | --- | --- | --- | --- | --- | --- | --- | --- | --- | --- |
| 1. Trisomy Blood | 33.55 | 32.41 |  |  |  |  |  |  |  |  |  |  |  |  |  |  |
| 2. Trisomy Buccal | 46.93 | 25.44 | .82** |  |  |  |  |  |  |  |  |  |  |  |  |  |
| 3. Proband IQ | 3.68 | 0.84 | -.28 | -.50* |  |  |  |  |  |  |  |  |  |  |  |  |
| 4. Proband GDS | 13.63 | 6.22 | -.14 | .03 | -.16 |  |  |  |  |  |  |  |  |  |  |  |
| 5. Proband GAS Total | 19.76 | 8.84 | -.04 | .24 | -.07 | .80** |  |  |  |  |  |  |  |  |  |  |
| 6. Proband GAS Worries | 9.35 | 3.74 | -.24 | .02 | -.08 | .75** | .86** |  |  |  |  |  |  |  |  |  |
| 7. Proband GAS Fears | 4.90 | 3.63 | .08 | .43** | -.04 | .53** | .75** | .41** |  |  |  |  |  |  |  |  |
| 8. Proband GAS Physical | 5.47 | 3.37 | .05 | .14 | .10 | .71** | .85** | .69** | .47** |  |  |  |  |  |  |  |
| 9. Parent GDS | 7.55 | 4.61 | .23 | .07 | -.08 | .45** | .33* | .27 | .22 | .41** |  |  |  |  |  |  |
| 10. Parent GAS Total | 13.60 | 7.12 | .13 | .11 | .21 | .45** | .49** | .33* | .41** | .46** | .60** |  |  |  |  |  |
| 11. Parent GAS Worries | 6.43 | 3.51 | .10 | -.00 | .20 | .31 | .30 | .21 | .18 | .31* | .51** | .89** |  |  |  |  |
| 12. Parent GAS Fears | 4.40 | 2.75 | .06 | .24 | .14 | .38* | .52** | .25 | .60** | .45** | .43** | .74** | .42** |  |  |  |
| 13. Parent GAS Physical | 2.87 | 2.31 | .17 | .05 | .09 | .51** | .48** | .43** | .33* | .43** | .70** | .85** | .73** | .45** |  |  |
| 14. Age | 23.52 | 9.68 | -.22 | -.04 | -.01 | -.26 | -.10 | .07 | -.25 | -.17 | -.21 | -.24 | -.28* | -.10 | -.19 |  |
| 15. Gender (Male) | 23 | 37.1% | .28* | .28 | -.10 | .27 | .22 | .16 | .12 | .31* | .34* | .26 | .29* | .14 | .23 | -.09 |

*Note.* *M* and *SD* are used to represent mean and standard deviation, respectively.
IQ = parent-reported estimated IQ bands (e.g. severe intellectual disability (IQ < 49), moderate intellectual disability (IQ = 50-69), mild intellectual disability (IQ = 70-79), average (IQ = 80-109), above average (IQ > 110)). IQ bands were coded such that higher scores reflected higher estimated IQ, GAS = Glasgow Anxiety Scale, GDS = Glasgow Depression Scale.
\* indicates $p < .05$ . \*\* indicates $p < .01$ .

Thirty-eight (79%) probands reported symptoms at or above the recommended cutoffs for either the GDS or the GAS.

### Correspondence between Proband and Parent report

Overall, the pattern of correlations within proband or parent, and between proband and parent demonstrated both inter-rater and intra-rater reliability, supporting the construct validity of the assessments (Table 2). We found evidence of higher correlations within rater compared to between parent and proband. The correlation between proband-reported GAS Total and GDS was r = .80, and between the parent-reported GAS Total and GDS was r = .60. Correlations between parent and proband on the same construct were mostly positive and significant (*r*s = .45, .49, .21, .60, .43) for GDS, GAS Total, GAS Worries, GAS, Fears, and GAS Physical, respectively.

### Tissue-Specific Associations with Percentage of Trisomy 21

Correlations between percent trisomy in blood and buccal specimens with measures of internalizing symptoms are reported in Table 2. The only significant association between internalizing symptoms and percent trisomy was found for the correlation between buccal trisomy and proband reported specific fears subscale on the GAS-ID (r = .43, p = .007), which indicated that 18.4% of the variance in specific fears is associated with the percentage of trisomic cells. The correlation between buccal cell trisomy and parent-reported specific fears on the GAS-CS was in the same direction and was the strongest of all parent-reported outcomes. However, it did not reach statistical significance (r = .24, p = .15).

The results of the linear multiple regressions, predicting regression internalizing problems on trisomy percentage, age, and gender are found in Tables 3 and 4. The association between buccal-derived trisomy percentage and proband-reported fear in the linear regression remained significant after controlling for age (centered at age 12) and gender (beta = 0.05, t(33) = 2.43, p = 0.020; Std. beta = 0.40). However, the overall model was not statistically significant and yielded a moderate proportion of variance (R2 = 0.20, F(3, 33) = 2.73, p = 0.059, adj. R2 = 0.13).

**Table 3.** Linear Multiple Regression of Percent Trisomy from Buccal on Proband- and Parent-Reported Internalizing Symptoms.

|  | Proband-Report |  |  |  |  | Parent-Report |  |  |  |  |
| --- | --- | --- | --- | --- | --- | --- | --- | --- | --- | --- |
|  | Depression | Total Anxiety | Fears | Worries | Physical | Depression | Total Anxiety | Fears | Worries | Physical |
| (Intercept) | 10.77 ±<br>(1.81) | 16.29 ±<br>(2.50) | 4.47 ±<br>(0.96) | 8.36 ±<br>(1.12) | 3.81 ±<br>(0.93) | 5.93 ±<br>(1.35) | 11.38 ±<br>(2.07) | 4.08 ±<br>(0.74) | 4.86 ±<br>(1.02) | 2.44 ±<br>(0.73) |
| Trisomy Buccal | -0.48<br>(1.03) | 1.44<br>(1.47) | 1.37 *<br>(0.56) | -0.09<br>(0.66) | 0.18<br>(0.55) | -0.06<br>(0.84) | 0.09<br>(1.30) | 0.49<br>(0.46) | -0.38<br>(0.64) | -0.01<br>(0.46) |
| Age Centered | -0.87<br>(1.00) | 0.29<br>(1.42) | -0.12<br>(0.54) | 0.27<br>(0.64) | -0.00<br>(0.53) | -0.07<br>(0.84) | 0.78<br>(1.26) | 0.75<br>(0.45) | -0.05<br>(0.62) | 0.08<br>(0.44) |
| Gender (Female) | 4.37<br>(2.21) | 4.74<br>(3.08) | 0.78<br>(1.18) | 1.50<br>(1.39) | 2.20<br>(1.15) | 2.74<br>(1.77) | 3.80<br>(2.68) | 0.37<br>(0.96) | 2.53<br>(1.32) | 0.90<br>(0.94) |
| N | 36 | 37 | 37 | 36 | 36 | 34 | 35 | 35 | 35 | 35 |
| R2 | 0.14 | 0.12 | 0.20 | 0.04 | 0.12 | 0.09 | 0.07 | 0.14 | 0.11 | 0.03 |

All continuous predictors are mean-centered and scaled by 1 standard deviation.
± p < 0.001; \*\* p < 0.01; \* p < 0.05.

**Table 4.** Linear Multiple Regression of Percent Trisomy from Blood on Proband- and Parent-Reported Internalizing Symptoms.

|  | Proband-Report |  |  |  |  | Parent-Report |  |  |  |  |
| --- | --- | --- | --- | --- | --- | --- | --- | --- | --- | --- |
|  | Depression | Total Anxiety | Fears | Worries | Physical | Depression | Total Anxiety | Fears | Worries | Physical |
| (Intercept) | 10.69 ±<br>(1.80) | 15.32 ±<br>(2.32) | 3.59 ±<br>(0.85) | 8.26 ±<br>(1.01) | 3.96 ±<br>(0.90) | 6.45 ±<br>(1.18) | 11.78 ±<br>(1.84) | 3.83 ±<br>(0.72) | 5.49 ±<br>(0.86) | 2.45 ±<br>(0.64) |
| Trisomy Blood | -1.92<br>(1.06) | -1.08<br>(1.41) | -0.06<br>(0.52) | -0.90<br>(0.62) | -0.11<br>(0.55) | 0.64<br>(0.78) | 0.23<br>(1.19) | 0.07<br>(0.46) | -0.08<br>(0.56) | 0.24<br>(0.41) |
| Age Centered | -1.72<br>(1.02) | -0.12<br>(1.35) | -0.47<br>(0.50) | 0.41<br>(0.60) | -0.06<br>(0.53) | -0.22<br>(0.76) | -0.62<br>(1.12) | 0.06<br>(0.44) | -0.48<br>(0.53) | -0.20<br>(0.39) |
| Gender (Female) | 3.63<br>(2.19) | 4.72<br>(2.87) | 1.16<br>(1.06) | 1.22<br>(1.26) | 1.83<br>(1.11) | 2.00<br>(1.57) | 3.60<br>(2.42) | 0.60<br>(0.94) | 2.11<br>(1.13) | 0.89<br>(0.84) |
| N | 39 | 40 | 40 | 39 | 39 | 40 | 41 | 41 | 41 | 41 |
| R2 | 0.16 | 0.07 | 0.06 | 0.09 | 0.07 | 0.10 | 0.09 | 0.01 | 0.12 | 0.07 |

All continuous predictors are mean-centered and scaled by 1 standard deviation.
± p < 0.001; \*\* p < 0.01; \* p < 0.05.

## Discussion

This is the first study to investigate psychiatric phenotypes among people with mDS. We found evidence from both parent and self-report screening measures that individuals with elevated levels of depression (53%) and anxiety (76%) symptoms are common in people with mDS. Although these numbers do not represent diagnostic criteria, these findings are consistent with several study results finding elevated rates of depression and anxiety disorders in people from the larger Trisomy 21 population (e.g. mood disorders ranging from 5.0% - 15%, anxiety disorders ranging from 7 – 30% (Baumer et al., 2023; Collacott et al., 1992; Mantry et al., 2007; Myers & Pueschel, 1991; Patti & Tsiouris, 2006; Raffaele et al., 2022; Rivelli et al., 2022; Vicari et al., 2016). The fact that the number of people scoring above the clinical cut-offs in this sample is substantially higher than reported diagnosed cases in other studies is to be expected. Screening instruments often result in higher levels of predicted cases compared to diagnostic criteria primarily because these instruments are designed to be highly sensitive. They aim to identify as many potential cases as possible, including mild or borderline cases, to ensure that individuals who may need further evaluation or treatment are not overlooked. Consequently, these tools may overestimate the prevalence of depression and anxiety by including individuals who do not meet the full, strict diagnostic criteria used in clinical settings. This overestimation is a trade-off for the benefit of not missing individuals who could benefit from further assessment and possible intervention.

The severity of internalizing symptoms among the sample of people with mDS varied according to the assessment instrument and informant; however, average severity scores did not differ by method of assessment (i.e., in person versus online). Symptom score values was generally higher with the self-report versus parent-report scales, which is consistent with the literature on child-parent discrepancies for internalizing symptoms (De Los Reyes & Kazdin, 2005). Much of the research on parent-child discrepancies of child mental health symptoms finds that children are more likely to report higher internalizing symptoms (e.g., anxiety, depression) than their parents. This finding is often attributed to the nature of internalizing symptoms, which are often less observable than externalizing symptoms, such as aggression or defiance.

The correlations between parent and proband reported symptoms on the same construct (21 - .60) was within range, and higher on average, compared to a meta-analysis on parent-child discrepancies for internalizing symptoms (mean *r* = .25;(De Los Reyes & Kazdin, 2005)). Parents usually completed their assessments before their proband, although in some cases due to COVID-data collection, parents assisted their children in completing the questions before answering their own. Together, these results also suggest that individuals with mDS, as with others with mild to moderate ID, are capable of reporting on their symptoms of depression and anxiety (Douma et al., 2006; Lindsay et al., 1994; Sánchez-Teruel & Robles-Bello, 2020), and that self-report measures should be incorporated into clinical practice, alongside informant report measures (Mileviciute & Hartley, 2015; Moss et al., 1996; Scheirs et al., 2023).

The overall association between percent trisomy and internalizing symptoms was weak. We found evidence of small to moderate effect sizes for the correlation between specific fears and percent trisomy from buccal cells, with moderate, and statistically significant, effects being observed for proband-reported fears. However, this relationship was not statistically significant for parent-reported fears. This finding is suggestive of a potential relationship between “dosage” effects for trisomy 21 and a predisposition towards fearfulness; however, this effect was not corrected for multiple testing and more research with a larger sample size is needed to confirm this finding.

The lack of an association between IQ band and fear (or other internalizing symptoms) suggests that this effect is unlikely to be accounted for by general cognitive ability. This is notable given that trisomy is associated with general cognitive ability. The finding of an association between fear and percent trisomic cells in buccal specimens, but not blood, is consistent with the results of previous studies that found associations between other cognitive phenotypes (i.e., IQ) and the percentage of trisomic cells in buccal smear, but not blood, specimens (Papavassiliou et al., 2009). Our study also replicated the previous finding in which parent-reported IQ-bands were significantly associated with buccal cell trisomic values, but not blood trisomic values. Although the samples were partially overlapping, they represent unique IQ data collection.

## Limitations

The results of this study should be considered within the context of several limitations. First, although the sample size is large for studies of people with mDS, which is an uncommon condition, the available sample size for assessing the trisomic cell analyses (n = 35 – 41), was relatively small (in part, due to COVID-19 restrictions). Thus, we were only powered to detect medium to large effects. Additionally, the sample may not be representative of the general population of people with mDS or DS. The prevalence of DS is largely consistent with racial and ethnic distribution of the overall population (Canfield et al., 2014). We attempted to balance the convenience sample of families attending a research retreat for families with mDS, who may be more actively engaged in research and have greater resources for travel, by recruiting through DS advocacy organizations and clinics across the country, traveling to participants, and recruiting through the DS-CONNECT registry, (which contributed to the subset of probands who did not have trisomy values). Unfortunately, engagement of minoritized racial and ethnic groups in advocacy organizations and research registries is also lacking, and the composition of our sample also reflects this limitation. Intersectionality in race, gender, and disability have been shown to be associated with a wide range of negative health consequences (Erevelles & Minear, 2010; Hassiotis, 2020), including among people with DS (Centers for Disease Control and Prevention (CDC), 2001). It is critical for future work to more intentionally engage underrepresented populations in research in order to address these disparities.

This study relied on quantitative measures of internalizing symptoms rather than clinical diagnosis. However, many have argued that psychiatric phenotypes are best represented as quantitative rather than qualitative constructs (Plomin et al., 2009; Waszczuk et al., 2020). We were also not able to formally assess IQ, language abilities, and other potential comorbid medical and cognitive conditions that may account for at least a portion of the observed patterns in internalizing symptoms. More research is needed to determine if the high rates of internalizing symptoms observed in this sample are related to other traits associated with trisomy 21 (e.g., sleep apnea; (G. Capone et al., 2006; G. T. Capone et al., 2011, 2013). Our group is currently conducting a larger scale study to address these limitations.

## Implications

Our study reveals that individuals with mDS, like people with non-mosaic DS, have a high risk for depression and anxiety symptoms, emphasizing the need for routine mental health screening and intervention in clinical settings, regardless of the percentage of trisomy detected in their cells. In light of the recent United States Preventative Task Force recommendations for routine screening of anxiety and depression in the general child (US Preventive Services Task Force, Mangione, Barry, Nicholson, Cabana, Chelmow, et al., 2022; US Preventive Services Task Force, Mangione, Barry, Nicholson, Cabana, Coker, et al., 2022) and adult population (US Preventive Services Task Force, 2023b, 2023a), these findings indicate that people with mDS, and likely non-mosaic DS, should not be excluded from screening. We also found good internal consistency and evidence of convergent validity of the Glasgow Depression (Cuthill et al., 2003) and Glasgow Anxiety (Mindham & Espie, 2003) self-report scales for people with mDS, supporting the use of self-report scales whenever possible for this population, in both research and clinical contexts. Early identification and treatment can improve quality of life and reduce healthcare costs through early intervention and treatment.

The research implications of our findings include exploring possible underlying mechanisms contributing to increased depression and anxiety risk, examining intervention effectiveness, and conducting longitudinal studies to understand symptom progression and associations with other health outcomes over time. Investigating the impact of these symptoms on functioning, such as social relationships and employment, can guide intervention development and support services.

Finally, our findings have public policy implications, including the need for accessible mental health services and provider training for individuals with rare genetic conditions such as mDS, as well as increased awareness and understanding among policymakers, healthcare providers, and the public. Adequate funding for research on rare genetic conditions can improve knowledge of biological and psychological factors contributing to observed behavioral phenotypes, leading to targeted interventions and support services for this underserved population.

## Acknowledgements

With gratitude, the authors wish to acknowledge the contribution of DS-Connect® (The Down Syndrome Registry) which is supported by the *Eunice Kennedy Shriver* National Institute of Child Health and Human Development (NICHD), National Institutes of Health (NIH), for study recruitment, and all the wonderful individuals and family members who participated in this study. We also wish to acknowledge the contribution of Dr. Steve Aggen for mentorship and guidance on data analytics throughout the project. Several students and trainees contributed to this project through data collection and data entry including Johnnie Mortensen and Rachel Siefring.

## Ethics Approval

All participants provided informed consent and/or parental permission with assent, as appropriate based on age and capacity to consent. All study procedures were reviewed and approved by the Institutional Review Board of Virginia Commonwealth University.

## Data availability

Due to the risk of reidentification, the data will not be posted publicly. However, analytic scripts and limited de-identified data may be made available by request from the corresponding author.

